# The Cost of Living Index as a Primary Driver of Homelessness: A Cross-State Analysis

**DOI:** 10.1101/2023.09.19.23295803

**Authors:** Thomas F. Heston

## Abstract

**Background:** Homelessness persists as a critical global issue despite myriad interventions. This study analyzed state-level differences in homelessness rates across the United States to identify influential societal factors to help guide resource prioritization.

**Methods:** Homelessness rates for 50 states and Washington D.C. were compared using the most recent data from 2020-2023. Twenty-five variables representing potential socioeconomic and health contributors were examined. Given non-normal distributions, nonparametric statistical techniques, including correlation and predictive modeling, identified significant factors.

**Results:** The cost of living index, mainly influenced by housing, transportation, and grocery costs, showed the strongest positive correlation with homelessness rates (all p <0.001). Unemployment, alcohol binging, taxes, and poverty were also influential factors. Opioid prescription rates demonstrated an unexpected negative correlation. Random forest classification emphasized the cost of living index as the primary contributor, with housing costs presenting the largest influence.

**Conclusion:** This state-level analysis revealed the cost of living index, predominantly driven by housing expenses, as the foremost factor associated with homelessness rates, greatly outweighing other variables. These findings can help inform resource allocation to mitigate homelessness through targeted interventions.

## Introduction

Homelessness remains a pervasive global issue. In the United States, over 500,000 people were estimated to be without adequate housing on any given night in 2020. Europe reported even higher figures, exceeding 700,000. In these regions, the estimated per capita rate of homelessness is approximately 0.2%, equating to one in every 500 individuals (Henry et al., 2021). Additionally, mobile populations like people seeking refuge and migrants, who comprise about 3% to 5% of the global populace, confront similar challenges (Crawford et al., 2022). These alarming statistics emphasize the urgent need for efficacious and targeted measures to alleviate homelessness.

One of the major issues contributing to homelessness is the rising cost of housing. Housing accounted for a fifth of inflation in 2022 in the United States. However, by March 2023, the housing inflation rate rose to 2.6 percentage points, accounting for half of the annual consumer price index inflation (White House Staff, 2023). With rental prices rising, even minor improvements in housing affordability are estimated to substantially reduce the incidence of homelessness (Quigley & Raphael, 2001). Additionally, job losses and stagnant wages can further exacerbate the problem, making it difficult for people to secure and maintain housing. These economic conditions affecting housing affordability significantly impact homelessness.

The health consequences of homelessness are severe. Estimates suggest a lifespan reduction of 5 to 10 years for homeless individuals (Funk et al., 2022). Furthermore, age-adjusted mortality rates for the homeless in New York shelters are two to four times higher than the general population (Barrow et al., 1999). Homeless children have elevated developmental delays and abuse rates but reduced access to social services to address these issues (Vostanis et al., 1997).

Various intervention strategies have yielded some success. When augmented with social services, permanent supportive housing improves long-term stability (Aubry et al., 2020). While income-based interventions enhance housing stability, they have not been shown to affect psychiatric symptoms, substance abuse, or employment outcomes. Offering immediate housing without preconditions such as sobriety has demonstrated mixed results. While effective in achieving initial housing stability, participants are more prone to incarceration and are less prepared for independent living upon discharge (Hall et al., 2020). A study in Ireland revealed that almost 90% of participants maintained their initial tenancy, indicative of low housing turnover (Greenwood, Byrne & O’Shaughnessy, 2022). However, these programs show uneven success in homeless youth, although it is notably effective for those with mental health conditions (Kozloff et al., 2016).

Despite these programmatic successes, homelessness rates have risen, underscoring the need for more efficacious interventions. This study aims to identify societal-level factors influencing homelessness by examining variations in rates across all 50 U.S. states and Washington, D.C. The insights garnered will help tailor community-specific solutions, enhancing the effectiveness of homelessness intervention programs.

## Materials and Methods

### Sample

State-level data were collected for the 50 US states and Washington, DC. The most recent data available as of September 2023 were obtained. Variables examined included homelessness, unemployment, cost of living index, grocery cost index, housing cost index, utility cost index, transportation cost index, health cost index, poverty rate, per capita real GDP, drug overdose mortality, median household income, incarceration rate, gasoline price, Gini coefficient, average state and local taxes, percentage of income spent on housing (renters), binge drinking prevalence, opioid prescriptions per capita, smoking prevalence, high school graduation rate, cigarette tax, alcohol consumption per capita, 2020 presidential election results, state population, incarceration rate, and sanctuary status. When possible, data were retrieved from official government sources. Ethical review board approval was not required as this study analyzed existing public data. No human or animal research was performed. Data collected for this analysis is publicly available in the Zenodo repository (Heston, 2023).

### Statistical Analysis

Both conventional statistical tests and machine learning techniques were utilized to examine associations with homelessness. *IBM SPSS Statistics* (version 29) was utilized for all analyses except for *random forest*, which was done using *Python*. Normality was assessed using the *Shapiro-Wilks* test. *Spearman correlation* with *Bonferroni correction* evaluated associations between variables. Machine learning techniques, including *exhaustive CHAID, classification and regression trees* (CRT), and *random forest regression*, were utilized to identify variables associated with homelessness rates. *Exhaustive CHAID* is a segmentation modeling approach that identifies groups using chi-square tests. *CRT* builds decision trees for classification or regression objectives using recursive binary splitting. *Random forest* is an ensemble technique aggregating results from many decision trees built using random subsets of variables and samples. These were followed by *automatic linear modeling* and *backward linear regression*, manually removing nonsignificant variables. *Collinearity tolerances* were evaluated for multicollinearity, and autocorrelation was evaluated with *Durbin-Watson* testing. A *test of proportions* compared binary categorical variables (political party and sanctuary status) with homelessness.

## Results

The data were not normally distributed per Shapiro-Wilks testing (p < 0.05). Transformation attempts were unsuccessful in normalizing distributions; thus, nonparametric tests were primarily utilized.

Significant correlations with homelessness rates were found for the housing cost index, cost of living index, transportation cost index, grocery cost index, cigarette excise tax, and opioid prescriptions per capita (all p < 0.001). Higher opioid prescription rates were associated with lower homelessness (Table 1).

**Table 1.** Spearman correlations with homelessness.

| Variable | Coefficient | p-value |
| --- | --- | --- |
| Housing Cost Index | 0.721 | < 0.001 |
| Cost of Living Index | 0.683 | < 0.001 |
| Transportation Cost Index | 0.628 | < 0.001 |
| Grocery Cost Index | 0.618 | < 0.001 |
| Cigarette Excise Tax | 0.499 | < 0.001 |
| Opioid Prescriptions per Capita | - 0.464 | < 0.001 |

Housing, transportation, and groceries were the key drivers of the cost of living index. Due to multicollinearity, these were consolidated into the overall cost of living index for the classification and regression modeling analyses.

Decision tree-based methods were then utilized to help further clarify and identify significant factors associated with homelessness. The *exhaustive CHAID* analysis was done with five parent and five child nodes, with the *Bonferroni correction* not enforced during model building. The resultant F values were normalized to add up to 100% to determine relative importance. This identified the cost of living index (0.642), state and local taxes (0.179), alcohol binging per capita (0.098), and opioid prescriptions per capita (0.081) as the most important factors, with a risk estimate of 47.8 and a standard error of 14.7.

*CRT* analysis was then done with manual weaning of factors to produce the strongest model based on its risk estimate, and the importance factors normalized to add up to 100%. Using this method, the factors identified as significant were the cost of living index (0.502), unemployment rate (0.362), and poverty (0.136).

A *random forest* analysis was then done. After a step-by-step process involving removing the least important factors, the best model consisted of the cost of living index (0.642 importance), alcohol binge rate (0.122), unemployment (0.091), taxes (0.085), and poverty (0.060). This model explained 65% of the variation in homelessness with an R-squared value of 0.655. Adding in the other factors did not improve upon this model. Notably, adding back in the opioid prescription rate did not improve the model but slightly decreased the R-squared value to 0.632.

*SPSS Automatic Linear Modeling* was then done using its built-in automatic weaning of insignificant factors. The resultant model was reviewed, and any remaining insignificant factors were removed. Using this method, the factors identified as significant were the cost of living index (importance 0.722, p < 0.001), unemployment (0.120, p=0.003), alcohol binge rate (0.089, p=0.011), and taxes (0.069, p=0.023). Finally, a *linear regression* model was done using backward processing and manual removal of low correlates. Importance was based on the standardized beta coefficient. The final model, after backward elimination, identified as significant the cost of living index (importance 0.564, p < 0.001), unemployment (0.245, p=0.004), and alcohol binge rate (0.192, p=0.018) with an adjusted R-squared value of 0.643. *Collinearity tolerances* were 0.871, 0.835, and 0.880, respectively, consistent with no significant multicollinearity. *Durbin-Watson* was 1.956, consistent with no autocorrelation. The adjusted R Square for the model was 0.643.

Significant factors contributing to homelessness identified by the various models are summarized along with the average overall importance in Table 2.

**Table 2.**
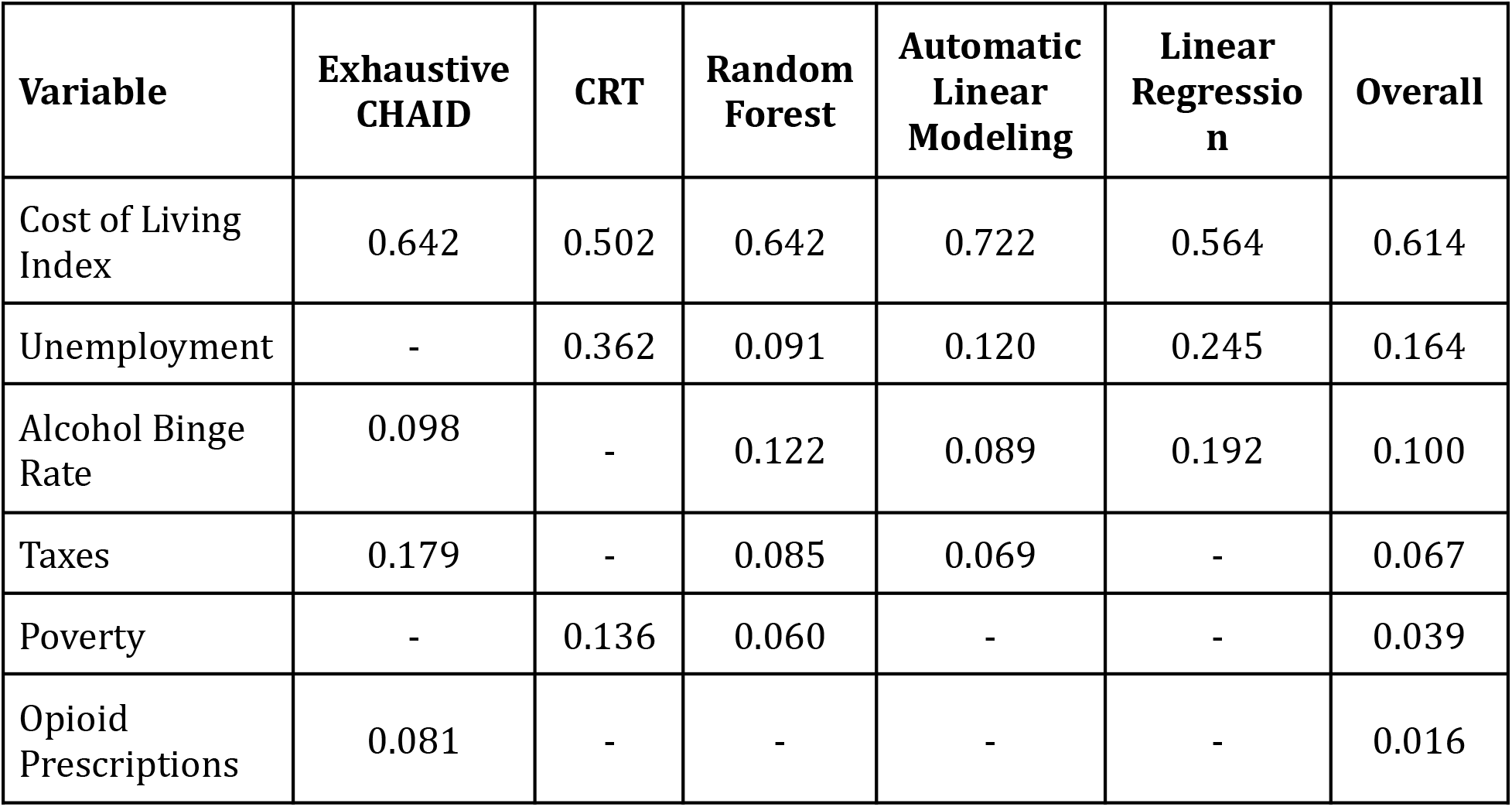
The relative importance of factors contributing to homelessness.

| Variable | Exhaustive CHAID | CRT | Random Forest | Automatic Linear Modeling | Linear Regression | Overall |
| --- | --- | --- | --- | --- | --- | --- |
| Cost of Living Index | 0.642 | 0.502 | 0.642 | 0.722 | 0.564 | 0.614 |
| Unemployment | - | 0.362 | 0.091 | 0.120 | 0.245 | 0.164 |
| Alcohol Binge Rate | 0.098 | - | 0.122 | 0.089 | 0.192 | 0.100 |
| Taxes | 0.179 | - | 0.085 | 0.069 | - | 0.067 |
| Poverty | - | 0.136 | 0.060 | - | - | 0.039 |
| Opioid Prescriptions | 0.081 | - | - | - | - | 0.016 |

The cost of living index was the most important factor in all five models and the most important overall, contributing 61.4% to the combined model. Unemployment was an important factor in four of the five models and overall contributed 16.4%. The alcohol binge rate was also identified as an important factor in four of the five models, with an overall contribution of 10%. Taxes were identified as important in three models: poverty in two and opioid prescription rate in one.

The cigarette excise tax was significantly correlated with homelessness by *Spearman correlation*. However, none of the three classification and two regression models identified it as a significant contributor.

## Discussion

This study identified the cost of living index, primarily driven by housing, transportation, and grocery costs, as the predominant factor associated with state-level homelessness rates. Across all models, the cost index was weighted 61.4% in importance as a contributing factor to homelessness. Unemployment, alcohol consumption, taxes, and poverty also emerged as significant contributors. Opioid prescription rates demonstrated an unexpected negative correlation. These findings highlight the multifactorial determinants of homelessness, with economic factors playing the predominant role.

The cost of living index was consistently the most influential variable. This aligns with previous research emphasizing the provision of low-cost housing and mental health services in mitigating homelessness (Elliot & Krivo, 1991). When this index increases, individuals face heightened housing instability and risk of homelessness. Housing, transportation, and groceries were key drivers. Our results suggest the need for interventions targeting these specific costs. Policies that enhance rental housing affordability, public transportation access, and nutritional assistance programs may help reduce homelessness.

Unemployment was also a major factor associated with homelessness rates, second only to the cost of living. Job creation, vocational training, and unemployment benefits likely aid in alleviating homelessness. The significance of alcohol consumption points to links between substance use disorders and housing instability that could be addressed through expanded mental health and addiction services.

Unexpectedly, higher opioid prescription rates correlated with lower homelessness, contrasting with previous studies (Cano & Oh, 2023). This warrants a deeper investigation into whether restrictive opioid policies are unintentionally displacing chronically ill patients toward dangerous street drugs and housing insecurity. Integrating harm reduction approaches into housing programs may mitigate overdose risks in this population.

This study possessed inherent limitations. The cross-sectional ecological design using group-level data restricted causal inference and omitted time-dependent effects. Though multiple analytical techniques were leveraged, the potential for unmeasured confounding variables remained. The reliance on secondary data introduced possible inaccuracies or biases. The generalizability of our models required context-specific interpretation given state-level heterogeneity.

Nonetheless, these findings can help guide resource allocation and policy decisions. Our results strongly endorse housing-first approaches that provide immediate, unconditional, permanent housing with ancillary services. Decreasing housing, transportation, and nutritional assistance costs should take priority. Expanding job creation and addiction treatment access may also reduce homelessness and secondary consequences like overdoses. A collaborative, multifaceted response will ultimately be essential to address this persistent public health crisis.

## Conclusion

This study identified the cost of living index, mainly housing, as the predominant factor associated with state-level homelessness rates, underscoring economic stability as a priority. Unemployment, substance use, and poverty also contributed significantly. Limitations like the ecological design restrict causal inference, yet the insights may aid future research and policy. The results strongly support housing-focused interventions and addressing income insecurity through job creation, transportation access, and nutritional assistance. Ultimately, collaborative, multifaceted efforts are needed to alleviate homelessness. This study highlights actionable targets to reduce homelessness and its associated health consequences.

## Data Availability

All data produced are available online at the Zenodo repository, doi: 10.5281/zenodo.8361377

https://zenodo.org/record/8361377

